# *PHEX* Gene Dosage Drives Meniere’s Disease and Related Audiovestibular Phenotypes in X-Linked Hypophosphatemia

**DOI:** 10.1101/2025.07.30.25332326

**Authors:** Paula Robles-Bolivar, David Bächinger, Arpan Bose, Kimberly Ramirez, Alison Brown, Amy F. Juliano, Jose Antonio Lopez-Escamez, Sharon G. Kujawa, Eva S. Liu, Sami S. Amr, Steven D. Rauch, Andreas H. Eckhard, Divya A. Chari

## Abstract

**Importance:** A subset of Meniere’s disease (MD) patients—defined by endolymphatic sac underdevelopment (ES hypoplasia), frequent bilateral disease, and strong male predominance—termed “MD-hp”, has emerged as a promising model for genetic investigation. We observed a striking enrichment of X-linked hypophosphatemia (XLH) caused by *PHEX* mutations among MD-hp patients, suggesting a shared genetic driver and immediate opportunities for biomarker discovery and targeted therapy development.

**Objective:** To test whether XLH and MD-hp share a causal, *PHEX* gene-driven relationship.

**Design:** Prospective, cross-sectional observational study.

**Setting:** Two tertiary academic centers.

**Participants:** Thirty-three adult XLH patients.

**Main Outcome Measures:** Pure-tone audiometry and speech-intelligibility testing; vestibular function via caloric and video head-impulse testing; symptom history fulfilling definite MD criteria; high-resolution CT assessment of ES hypoplasia (ATVA ≥ 140°) to define MD-hp; delayed 3D-FLAIR MRI detection of endolymphatic hydrops; and *PHEX*/pathway gene sequencing.

**Results:** Given population prevalences (XLH ≈ 0.005%; MD ≈ 0.2%; MD-hp ∼30% of MD), random co-occurrence would be ∼1 in 33 million. In our cohort, 6 of 33 met bilateral MD-hp criteria (XLH+MD-hp ≈ 18.2%; 1 in 5.5)—a > 6 million-fold enrichment—all hemizygous males (including two with fluctuating progressive sensorineural hearing loss (SNHL) but no vertigo). Two additional hemizygous males < 40 years of age displayed bilateral ES hypoplasia without clinical MD, and two males with mosaic or hypomorphic *PHEX* variants showed normal ES anatomy and no audiovestibular symptoms. No female carriers met MD-hp criteria; instead, five exhibited mild-to-moderate low-to-mid frequency sensorineural hearing loss without vertigo, and two had isolated conductive hearing loss.

**Conclusions and Relevance:** These findings support an inner ear–specific *PHEX* gene-dosage threshold model for MD-hp penetrance: complete loss-of-function in hemizygous males leads to bilateral ES hypoplasia and MD, whereas mosaic or partial loss variants in males—and heterozygosity in females—permit residual PHEX activity, resulting in milder or absent audiovestibular phenotypes. This genotype– endotype–phenotype linkage (complete PHEX loss → ES hypoplasia → MD) enables early risk stratification, personalized surveillance, and paves the way for targeted therapies in XLH patients.

## Introduction

Meniere’s disease (MD) is generally viewed as a syndrome in which diverse causes— autoimmune, viral, allergic, vascular, developmental, genetic, and others^1–8^—converge on shared pathophysiological pathways, producing the common clinical picture of episodic vertigo, fluctuating hearing loss, tinnitus, and aural fullness.^9^ Recent human histopathology^10–12^ and radiology studies^13–15^ have pinpointed the endolymphatic sac (ES) as the key site where these diverse insults presumably coalesce. In about 30% of patients, ES development arrests at a primitive, fetalLJlike stage—defining the “MDLJhp” endotype, characterized clinically (phenotype) by early disease onset (30s-40s), marked male predominance (>80%), frequent bilateral involvement (∼29%), and familial clustering of MD or hearing loss (∼41%), strongly suggesting a genetic contribution.^1,11^ In preliminary work, we noted several MDLJhp patients that were also affected by XLJlinked hypophosphatemia (XLH), a sex-linked dominant phosphate-wasting disorder caused by *PHEX* loss-of-function (LoF) mutations. XLH typically presents in childhood with impaired bone mineralization and growth, bone deformities, and dental abscesses;^16–19^ interestingly case reports and small series on adult XLH patients have documented associations with hearing loss and, occasionally, vestibular symptoms resembling MD.^20–22^

Here, we hypothesized that *PHEX* LoF variants underlying XLH predispose patients to the MD-hp endo-phenotype. To test this, we addressed the following questions: (1) Is the prevalence of MD-hp among XLH patients significantly higher than expected by chance? (2) If so, does MD-hp occur predominantly in hemizygous males, reflecting the X-linked inheritance of *PHEX*?, and (3) are certain *PHEX* variants associated with MD-hp penetrance? We recruited 33 XLH patients from two tertiary academic centers and performed comprehensive audiovestibular testing, high-resolution temporal bone CT for endolymphatic sac (ES) hypoplasia evaluation, and extended *PHEX* pathway sequencing, including bioinformatic modeling of variants of uncertain significance. We found an extraordinary enrichment of bilateral MD-hp—over six million times the expected rate—with cases confined to hemizygous males. Heterozygous females and males with mosaic or hypomorphic *PHEX* variants were either spared from MD-hp entirely or exhibited a milder endo-phenotype. These findings support a *PHEX* gene-dosage effect and unveil a complex genetic landscape governing MD-hp susceptibility.

## Methods

### Ethics

This study was approved by the IRBs of both academic institutions and adhered to institutional guidelines.

### Study Cohorts

XLH patients were recruited via Endocrinology Divisions at two tertiary academic centers using uniform inclusion criteria: adult age (≥18 years), XLH diagnosis per international consensus guidelines,^23^ using clinical, radiologic, and biochemical testing.^18^ Inclusion was independent of prior and current XLH treatment status (e.g., calcitriol, burosumab (anti–FGF-23 antibody)).^16,18,24^

### Audiometric Evaluation

Pure-tone air- and bone-conduction thresholds were measured in a sound-treated booth using an Interacoustics AC-40 audiometer (Interacoustics, Middlefart, Denmark). Air-conduction thresholds were assessed from 250 to 8000 Hz at interoctave intervals via TDH39P headphones (Telephonics, Santa Ana, CA, USA), and bone-conduction thresholds from 250 to 4000 Hz with a B71 vibrator (Radioear, Middlefart, Denmark) placed over the mastoid. The pure-tone average (PTA) was calculated at 500, 1000, and 2000 Hz. Speech recognition was measured with recorded CID W-22 word lists (Central Institute for the Deaf, St. Louis, MO, USA) presented with a contralateral masker at the level predicting maximal intelligibility and with a contralateral masker.

### Vestibular Function Testing

Bithermal caloric testing employed videonystagmography (Neuro Kinetics, Inc., Pittsburgh, PA, USA) using standard 30 °C and 44 °C irrigations via a Hortmann Aquamatic water stimulator (GN Otometrics, Taastrup, Denmark). Caloric paresis was quantified by Jongkee’s Index.^25^ The video head impulse test (vHIT) was conducted for all six semicircular canals with an ICS Impulse 3D unit (GN Otometrics, Taastrup, Denmark); head impulses (100–250°/s) were delivered until approximately ten valid impulses were obtained per canal, and vestibulo-ocular reflex (VOR) gain was calculated in OTOsuite (GN Otometrics). A VOR gain < 0.8 was considered abnormal.^26^

### Temporal Bone Imaging

Participants who consented underwent high-resolution CT (Discovery 750 HD, General Electric, Milwaukee, WI, USA; 0.6 mm slices with 0.2 mm overlap) or cone-beam CT (3D Accuitomo 170, Morita, Kyoto, Japan; 125 × 125 μm pixels, 0.5 mm slices). Axial reformats parallel to the lateral semicircular canal were exported to PACS. All participants with asymmetric hearing loss underwent gadolinium-enhanced MRI to exclude retro-labyrinthine pathology. Two participants additionally underwent delayed 3T gadolinium-enhanced MRI (750 GEM, GE Healthcare) with 3D-FLAIR sequences within four hours of contrast injection (0.7×0.7×0.6 mm voxels; 15 min acquisition) to grade endolymphatic hydrops.^27–31^

### Image analysis

On HRCT or CBCT images, the angular trajectory of the vestibular aqueduct (ATVA) was measured for each inner ear, using previously established methods.^11^ An ATVA with an angle α_exit_ ≥ 140° indicated ES hypoplasia, as previously defined.^10,11^ On delayed 3D-FLAIR MRI sequences, cochleo-vestibular hydrops was graded using the system reported by Bernaerts et al.^32^

### Clinical classification of hearing and balance symptoms

(1) Definitive MD was diagnosed according to established guidelines,^23^ with bilateral MD requiring criteria fulfillment in both ears; those meeting definitive MD criteria who also exhibited ATVA ≥ 140° (indicative of ES hypoplasia) were classified as MD-hp. In cases of long-standing MD without prior audiograms available, diagnosis was based on a study audiogram that had to show moderate-to-severe, “flat” (affecting mid-and low frequencies) sensorineural hearing loss (SNHL) combined with a history of episodic hours-long vertigo, and fluctuating auditory symptoms. (2) Patients with documented low-to-mid frequency SNHL—either fluctuating-progressive or solely progressive—without any vertigo or dizziness, were classified as fluctuating/progressive SNHL. (3) Those with isolated conductive hearing loss were classified as “CHL.”

### DNA extraction, library preparation, and sequencing

DNA extraction, library preparation, and sequencing were performed using two workflows depending on cohort. In the first cohort, buffy-coat DNA was extracted with Frozen FlexiGene and Blood DNA Finishing kits (Autogen, Holliston, MA, USA), quantified by PicoGreen (Thermo Fisher Scientific, Waltham, MA, USA), and prepared with the Illumina TruSeq DNA PCR-Free kit (Illumina, San Diego, CA, USA) on a Sciclone G3 workstation (PerkinElmer, Waltham, MA, USA), then sequenced (150 bp paired-end; ≥ 30× coverage) on a NovaSeq 6000 (Illumina) at MGB Personalized Medicine. In the second cohort, saliva DNA was collected via Oragene kits and prepIT® L2P (DNA Genotek, Ottawa, Canada), QC’d by NanoDrop 2000C (Thermo Fisher) and Qubit BR Assay (Thermo Fisher), exome-captured and enriched with Agilent SureSelectXT Human All Exon V6 (Agilent Technologies, Santa Clara, CA, USA), QC’d on a TapeStation D1000 (Agilent), and sequenced (≥ 100× coverage) on a NovaSeq 6000 by Macrogen (Seoul, South Korea).

### Bioinformatic preprocessing, variant filtering and prioritization pipeline

Read preprocessing, alignment to the GRCh38/hg38 reference genome, and variant calling were performed using the Illumina DRAGEN Germline Pipeline (v4.2.4) or GATK Best Practices (v4.5). Sequencing quality was assessed using standard metrics to ensure base call accuracy and sufficient coverage. SNVs and InDels were hard-filtered to remove low-confidence calls (exome: DP < 50, MQ < 20, GQ < 30, QD < 2; genome: GQ = 0, DP ≤ 1, QUAL < 3.0103). Variants were merged using BCFtools (v1.19) to generate a unified XLH variant dataset and annotated with Funcotator (v4.5.0.0) including functional (GenCode v34), clinical (ClinVar accesed March 2025), allele frequency (gnomAD v4.1) and computational predictors (alphaMissense, CADD v1.7). Our filtering pipeline then prioritized rare (allele frequency < 0.05) high- and moderate-impact variants in *PHEX*—specifically LoF and missense changes. For individuals lacking obvious *PHEX* hits, we extended our search to rare, high- or moderate-impact variants in *PHEX*-downstream genes, including all 23 members of the FGF23 signaling pathway (WikiPathways WP4790). Finally, we performed calling for large structural variants (LSV) using TIDDIT (v3.9.3) from CRAM files, followed by annotation with AnnotSV (v3.4.6). Resulting LSV were filtered to retain those in *PHEX* or *FGF23* pathway genes and inspected in IGV (v11). This tiered approach ensured comprehensive identification of both primary and modifier variants.^33^

### In silico structural and dynamics analysis of variants of uncertain significance

For variants whose consequences could not be clearly classified as LoF, we conducted in silico mutagenesis and three-dimensional structural modeling to assess potential pathogenicity. The predicted three-dimensional structure of the human wildtype *PHEX* protein was obtained from AlphaFold (Model ID: AF-P78562-F1-v4) and the amino acid substitutions were evaluated using DynaMut,^34^ estimating changes in Gibbs free energy (ΔΔG) and vibrational entropy (ΔΔSVib) to infer effects on protein stability and flexibility. PyMOL (Schrödinger, LLC, v3.1.5.1) was used to inspect and visualize local conformational changes and interatomic interactions introduced by the variant between wild-type and mutant models.

### Statistical analysis

A point-prevalence analysis was performed by calculating the expected co-occurrence of XLH and MDhp from published population prevalences and comparing it with the observed frequency in the study cohort. Age differences between males and females were assessed using Welch’s t-test. The association between PTA and age was evaluated via separate linear regression models for each indicated group, with correlation coefficients and p-values reported; p ≤ 0.05 was considered significant.

## Results

### High prevalence of audiovestibular phenotypes, particularly Meniere’s disease, in XLH patients

Among 33 XLH patients (Table 1), six out of ten males (#1–6) presented with bilateral low-frequency fluctuating SNHL; four of these (#1–4) also experienced episodic vertigo, aural fullness, and tinnitus, meeting criteria for bilateral definite MD (Figure 1A). In contrast, five females (#11–15) exhibited uni- or bilateral fluctuating/progressive SNHL without vertigo, and two (#16–17) had mild CHL (Figure 1A) despite normal tympanometry and radiological findings. Overall, males demonstrated more severe SNHL (Figures 1B) as compared to females (Figure 1C), word-recognition scores were similar across sexes—albeit with greater variability in males—and caloric responses and VOR gains were worse in males (Figure 1D). In symptomatic XLH patients of both sexes, PTAs exhibited an accelerated age-related increase—mirroring typical MD progression—while asymptomatic patients’ PTAs followed expected age-related norms (Figure 1E). These findings demonstrate a high prevalence of bilateral MD and related fluctuating/progressive SNHL phenotypes in XLH, disproportionately affecting males with greater severity.

**Figure 1.**
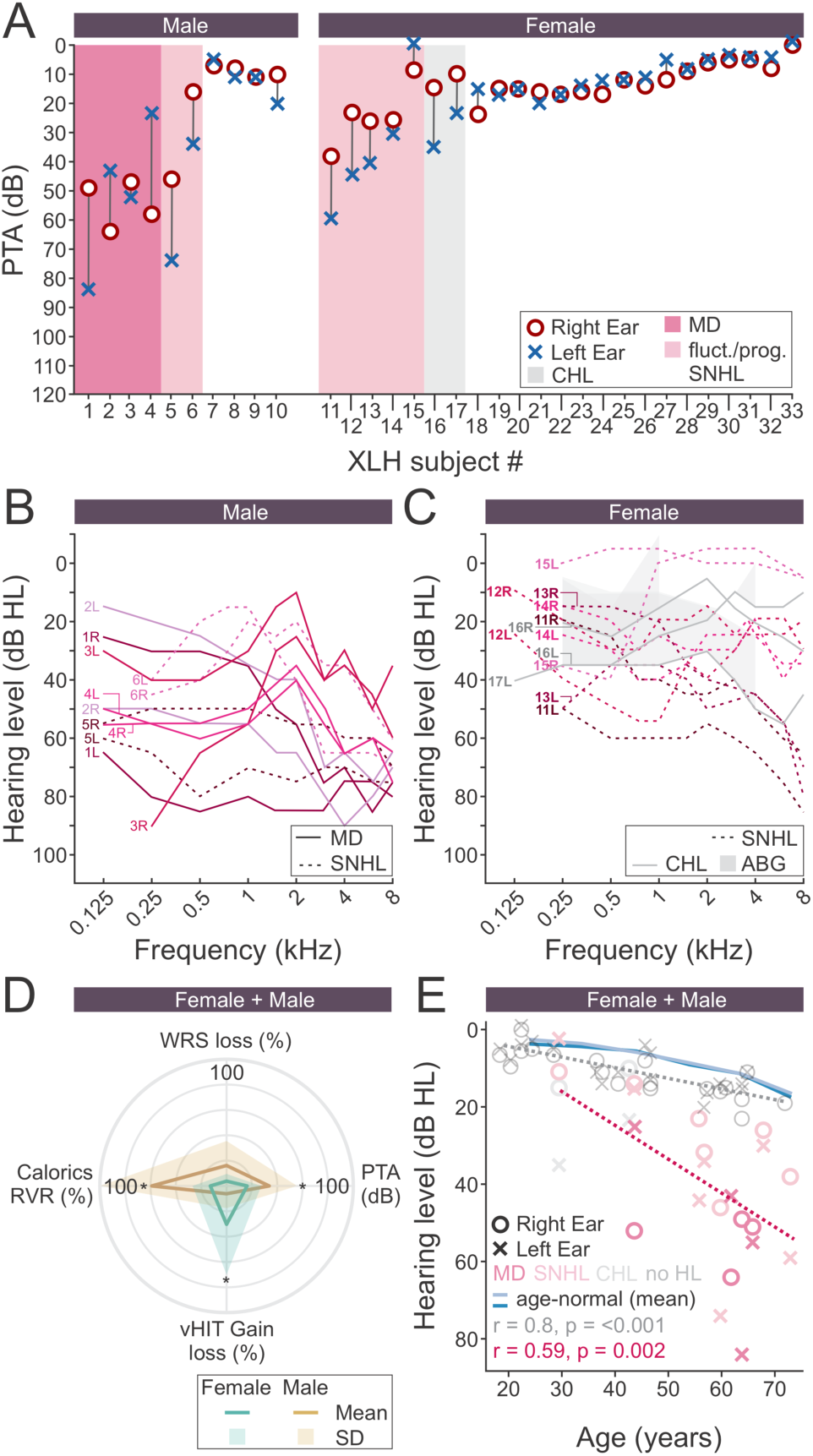
Audiovestibular profiles in XLH cohort. (**A**) Pure-tone averages (PTA) for all 33 subjects; shading indicates phenotype: bilateral MD (dark magenta), fluctuating/progressive SNHL (light magenta), CHL (gray). (**B, C**) Air-conduction thresholds from study audiograms for male (B) and female (C) patients with hearing loss; Air bone gap (ABG) for CHL shown as gray bands. (**D**) Mean ± SD comparison of males and females for four metrics: reduced vestibular (caloric) response (RVR), word recognition score (WRS) loss, PTA, and video head impulse test (vHIT) gain loss. (**E**) PTA versus age for all subjects, with separate regression lines for MD, fluctuating SNHL, CHL (magenta), and unaffected individuals (gray); the age-normalized PTA range^54^ is demonstrated by a light blue (females) and dark blue (males) lines.

**Table 1:**
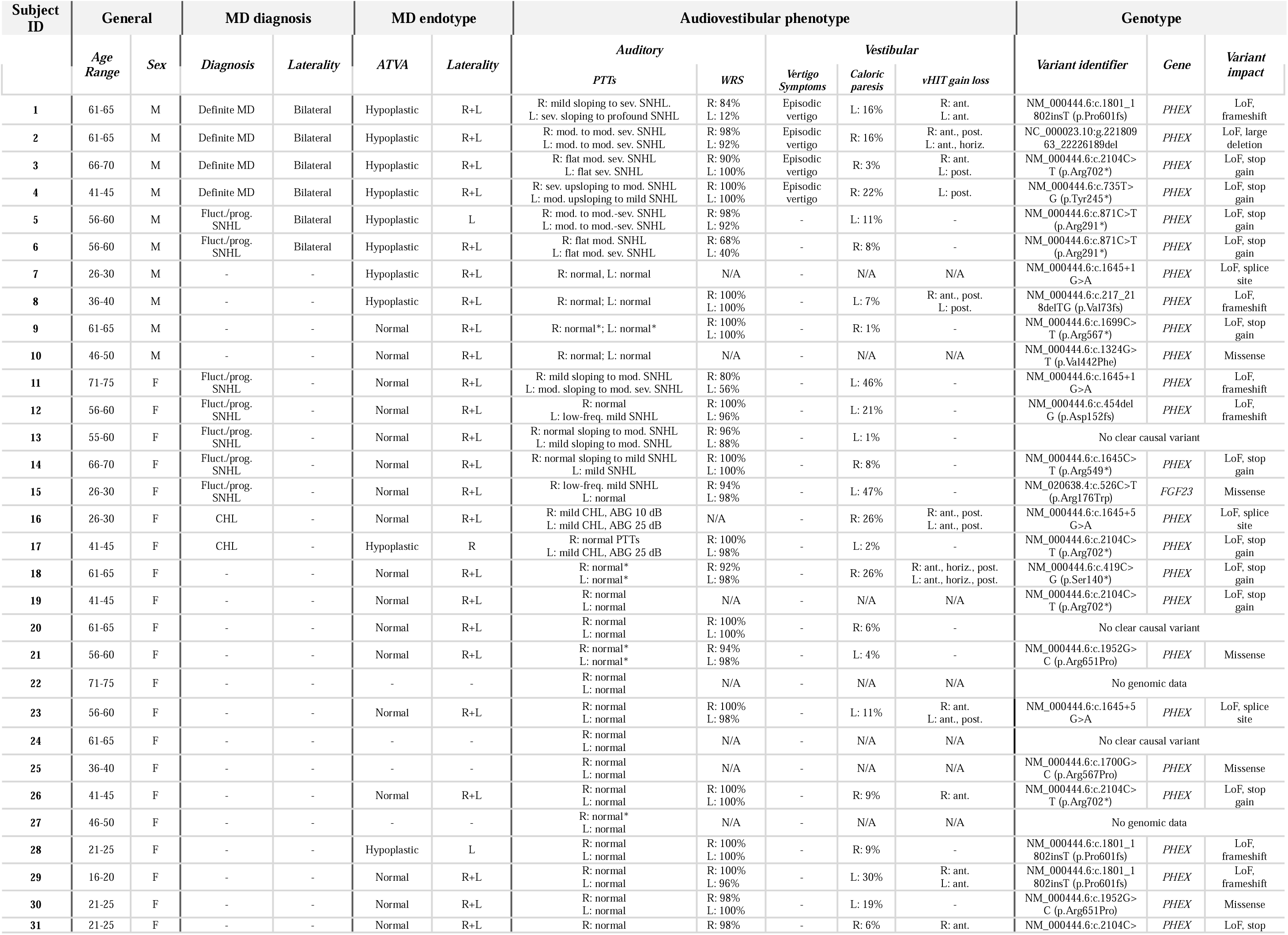
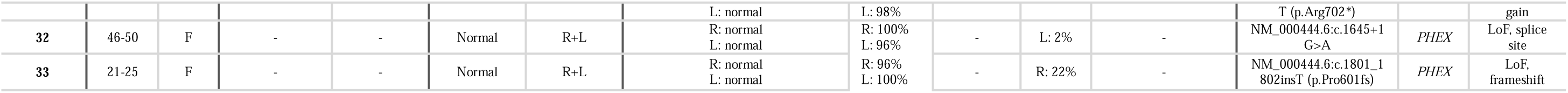
Characteristics of XLH subjects. . (*): age-adjusted pure tone thresholds. Abbreviations: PTT: pure tone threshold; WRS: word recognition score; R: right; L: left; ABG: air-bone gap; mod.: moderate; sev.: severe; ant.: anterior; post.: posterior; horiz.: horizontal; LoF: loss-of-function.

### Imaging reveals a male-predominant MD-hp endotype in XLH patients

Axial CT from an asymptomatic female XLH patient (ATVA=99°; Figure 2A–A″) and a male MD-hp patient (ATVA=164°; Figure 2B–B″) illustrates normal versus hypoplastic ES morphology, respectively. All six males with either definitive bilateral MD-hp (#1–4) or fluctuating/progressive SNHL without vertigo (#5–6) exhibited ATVA ≥ 140°, indicating a MD-hp endotype (Figure 2C). Two younger, asymptomatic males (#7, #8; < 40 years) also showed ATVA ≥ 140°, indicating preclinical ES hypoplasia likely to progress to MD.^13,35^ In contrast, two males (#9–10) and 20 females displayed ATVA < 140° bilaterally, while three females (#16, #28) had isolated unilateral ES hypoplasia.

**Figure 2.**
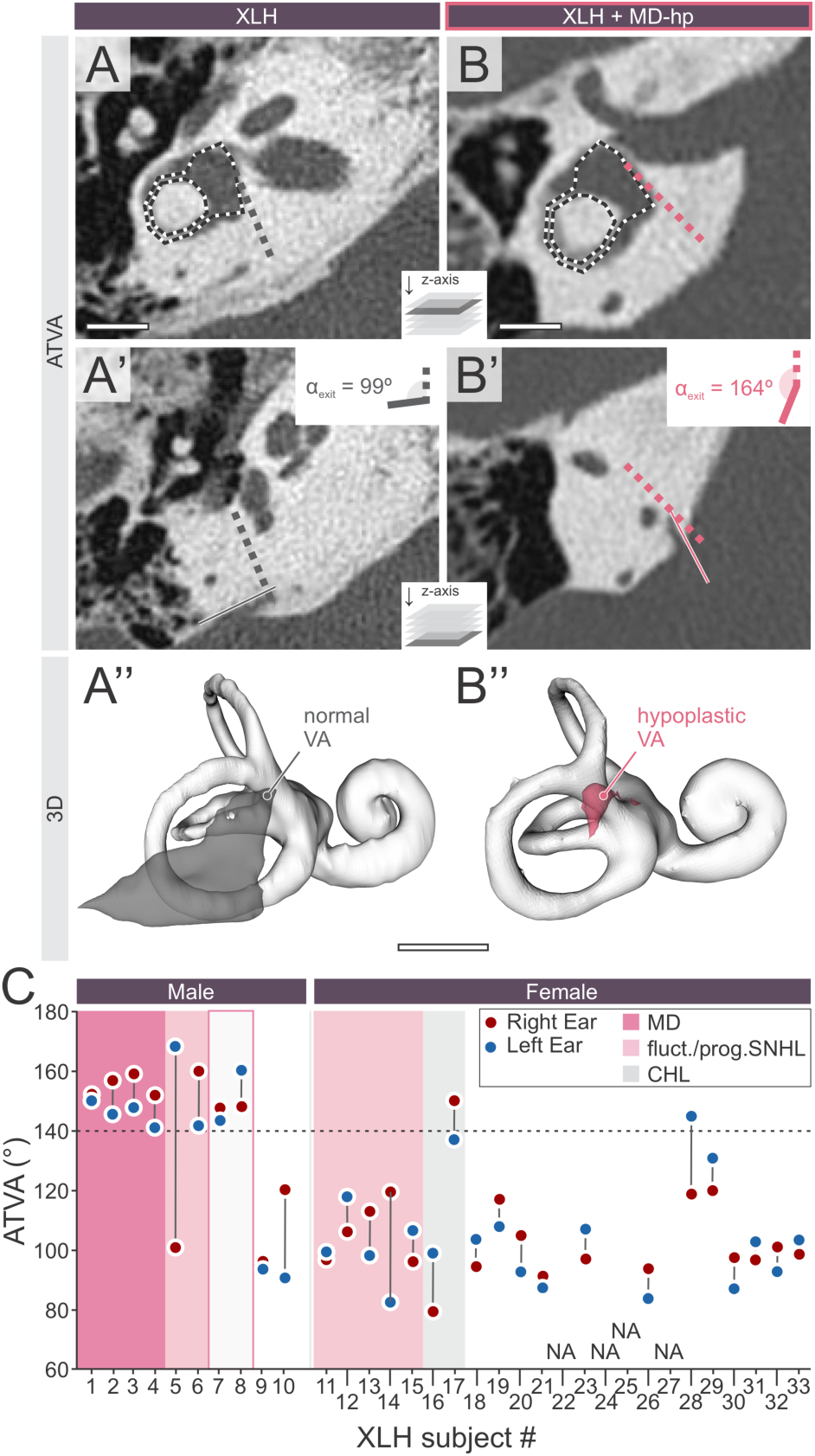
Example ATVA measurements and cohort ATVA distribution. (**A–A**″) Asymptomatic female XLH patient: axial CT with a template overlay on the vestibule and horizontal semicircular canal, showing the dotted proximal vestibular aqueduct (VA) trajectory and a second line along the distal VA; their intersection defines the α_exit angle (A′). A 3D reconstruction confirms a normal VA morphology (A″). (**B–B**″) Male XLH patient with MD-hp endo-phenotype: identical ATVA measurement on axial CT revealing an enlarged α_exit and ES hypoplasia (B′), with 3D reconstruction illustrating a hypoplastic VA morphology (B″). (**C**) α_exit angles (degrees) for all 33 XLH subjects (x-axis: subject ID; y-axis: α_exit), shaded by phenotype: bilateral MD (dark magenta), fluctuating/progressive SNHL (light magenta), and CHL (gray). Scale bars: 5 mm.

Delayed gadolinium-enhanced 3D-FLAIR MRI in two MD-hp males (#4–5) showed bilateral cochlear and vestibular hydrops (Figure 3A–B; Table 1), supporting their clinical bilateral MD diagnosis.^27,29,36^ A “halo-like” hypodensity around the otic capsule—seen in one MD-hp male(#1) and one female without SNHL (#28)—(Figure 3C–D; ^17,37^) may reflect XLH-related osteomalacia involving the otic capsule in these two patients, though its relevance to the audiovestibular phenotype remains uncertain. Together, these results define a male-predominant MD-hp endotype in XLH, characterized by ES hypoplasia and endolymphatic hydrops with high penetrance in hemizygous males.

**Figure 3:**
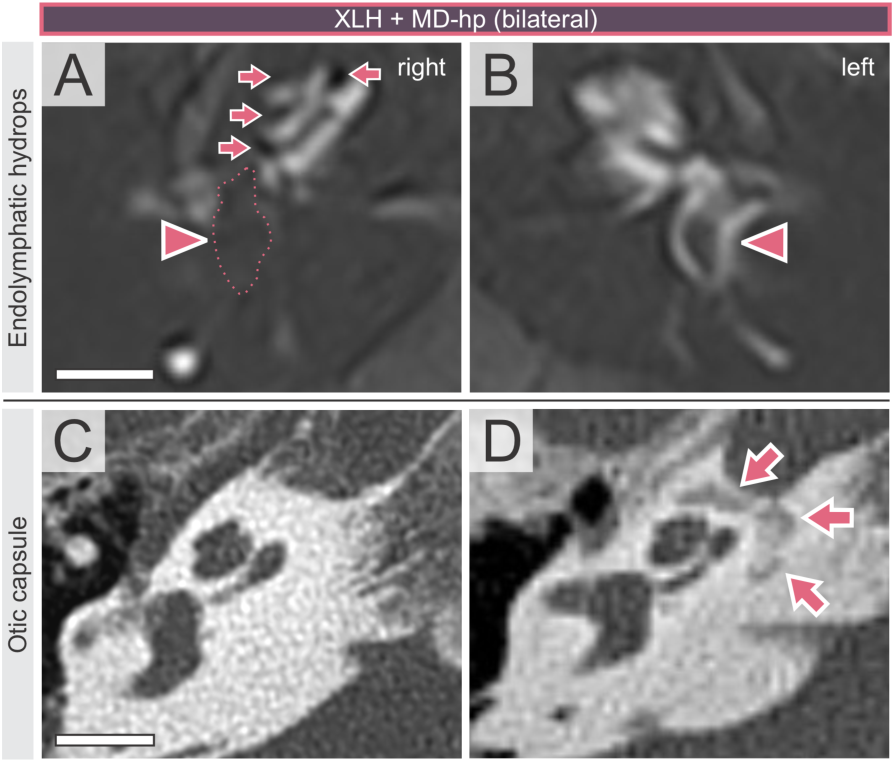
Endolymphatic hydrops and other imaging features in MD-hp associated with XLH. (**A, B**) Delayed gadolinium-enhanced 3D-FLAIR MRI of the right (A) and left (B) inner ears in a male XLH patient with MD-hp, highlighting endolymphatic hydrops (T2 signal devoid fluid spaces) in all cochlear turns (arrows) and the vestibule (arrow heads). (**C, D**) Axial CT of a XLH patient without (C) and with MD-hp (D); the latter exhibits a halo-like hypodensity surrounding the otic capsule, a feature also observed in other audiovestibular phenotypes. Scale bars: 5 mm.

### Epidemiological assessment shows prevalence of MD-hp in XLH far exceeds chance and is enriched in males

Using a standardized morbidity ratio, we compared the expected versus observed co-occurrence of MD-hp in XLH. Based on independent population prevalences (XLH ≈ 0.005%; MD ≈ 0.2%; MD-hp ≈ 30% of MD), a chance overlap would occur in only about 1 in 33.3 million individuals. Yet, among our 33 XLH patients, six hemizygous males (18.2%; ≈1 in 5.5) exhibited bilateral MD-hp—including two with ES hypoplasia and fluctuating/progressive SNHL without vertigo—amounting to an enrichment of over six million-fold. This dramatic enrichment strongly implicates *PHEX* deficiency as a causal driver of the MD-hp endophenotype.

### Spectrum of PHEX Variants and their correlation with MD-hp penetrance

Sequencing of *PHEX* and related pathway genes revealed that all hemizygous males with bilateral MD-hp (#1–4) and those with fluctuating/progressive SNHL plus ES hypoplasia (#5, 6) carried LoF *PHEX* variants (Table 2), as did the two asymptomatic young males with ES hypoplasia (#7, 8). No single *PHEX* variant was overrepresented among cases with MD-hp or fluctuating/progressive SNHL (Figure 4A). Among the males without MD-hp, one (#9) harbored a mosaic truncating *PHEX* mutation (allele frequency 93%; Figure 4B), and another (#10) carried a rare hemizygous missense substitution (p.Val442Phe) in *PHEX*’s catalytic domain. This variant is absent from gnomAD v4, predicted deleterious by in silico metrics (AlphaMissense 0.66; CADD 15.87), and structural modeling suggested steric clashes, modest destabilization (ΔΔG –1.17 kcal/mol), and reduced global flexibility(ΔΔSVib –0.88 kcal·molLJ¹·KLJ¹) (Figure 4C), consistent with a hypomorphic allele retaining residual protein function. Female XLH patients likewise carried heterozygous LoF and missense variants in *PHEX* and downstream *FGF23*, respectively—though three had no identifiable pathogenic variants—and no specific variant correlated with fluctuating/progressive SNHL or CHL, nor did any cluster within a specific protein domain (Figure 4A). Together, these data indicate that complete loss of *PHEX* activity in hemizygous males confers a high risk for MD-hp, whereas partial *PHEX* function—whether due to mosaicism, hypomorphic alleles in males, or heterozygosity in females—does not produce the MD-hp endo-phenotype. However, heterozygous females may still exhibit a milder, fluctuating-progressive SNHL or CHL phenotype, the basis of which remains to be determined.

**Figure 4.**
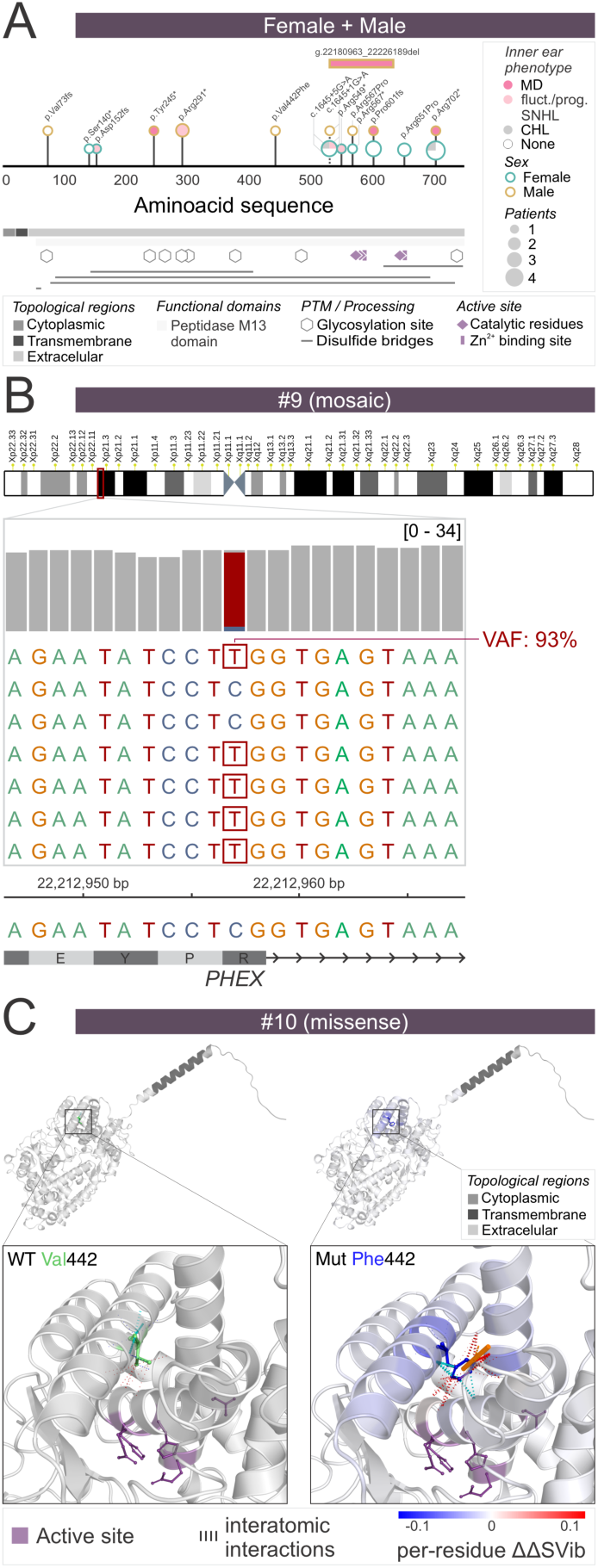
*PHEX* variant mapping and structural impact. (**A**) PHEX protein topology Tick marks indicate all study variants along the 749-amino-acid sequence. (**B**) X-chromosome schematic for subject #9 in the *PHEX* locus showing the truncating mosaic mutation with a 93 % allele fraction. (**C**) Overlay of predicted PHEX protein catalytic domain structures: wild-type (left) versus p.Val442Phe mutant (right), highlighting the appearance of steric clashes and local rigidification around the mutated residue.

**Table 2.** Causal variants in the XLH cohort. General information includes HGVS nomenclature, genomic coordinates (GRCh38), affected gene, variant type, predicted consequence and exon/intron number location. Prediction scores indicate *in silico* assessments of missense and splice site variants (e.g., AlphaMissense [αm], Combined Annotation Dependent Depletion [CADD], SpliceAI [SpAI] and Pangolin [Pang]); meanwhile Loss-of-function variants are marked as n.a. (not applicable) presumed clearly pathogenic. Allele frequencies (AF) are indicated for global and specific subpopulations including NFE (Non-Finnish European), AF/AMR (African/American), and AD/AMR (Admixed/American) from gnomAD v4.1 (aggregated data from genomes plus exomes). Clinical significance and ClinVar identifiers are provided for previously reported variants; otherwise, are indicated as *novel*. Significance with an asterisk (*) are based on variant described with the same predicted protein-level consequence, despite differing at the nucleotide/genomic change. LoF: Loss of Function; LSV: large structural variant; n.a.: not applicable; n.d.: not described; SNV: single nucleotide variant. Clinical and *in silico* predictors abbreviations: A: Ambiguous; I: indeterminate; LP: likely pathogenic, MB: moderate benign; MP: moderate pathogenic; P: pathogenic; SP: supporting pathogenic; US: uncertain significance; high SpAI and Pang scores indicate strong computational evidence for splicing impact.

| Impact | Variant identifier (HGVS) | Genomic coordinates | Gene | Variant Type | Consequence | Location | Prediction scores | gnomAD AF<br>global\|NFE\|AF\|AMR\|AD\|AMR | Clinical<br>significance<br>(ClinVar) | ClinVar ID | Subjects |
| --- | --- | --- | --- | --- | --- | --- | --- | --- | --- | --- | --- |
| High | NM_000444.6:c.2104C>T (p.Arg702*) | chrX 22245366 C/T | *PHEX* | SNV | LoF, stop gained | Exon 21/22 | n.a. | n.d. | P | 279872 | #3, #17, 19, 26, 31 |
|  | NM_000444.6:c.1801_1802insT (p.Pro601fs) | chrX 22221647 -/T | *PHEX* | short insertion | LoF, frameshift | Exon 18/22 | n.a. | n.d. | P* | novel | #1, 28, 29, 33 |
|  | NM_000444.6:c.1699C>T (p.Arg567*) | chrX 22212957 C/T | *PHEX* | SNV | LoF, stop gained & splice site | Exon 16/22 | n.a. | n.d. | P | 10822 | #9 |
|  | NM_000444.6:c.1645+1G>A | chrX 22190503 G/A | *PHEX* | SNV | LoF, splice site | Intron 15/21 | spAI: 0.98 (high); Pang: 0.78 (high); CADD: 34 (P) | 8.63E-07\| n.d.\| n.d.\| n.d. | P | 279871 | #7, 11, 32 |
|  | NM_000444.6:c.1645+5G>A | chrX 22190507 G/A | *PHEX* | SNV | LoF, splice-site | Intron 15/21 | spAI: 0.98 (high); Pang: 0.78 (high); CADD: 26.3 (SP) | n.d. | P | 951146 | #16, 23 |
|  | NM_000444.6:c.1645C>T (p.Arg549*) | chrX 22190502 C/T | *PHEX* | SNV | LoF, stop gained & splice site | Exon 15/22 | n.a. | n.d. | P | 279870 | #14 |
|  | NM_000444.6:c.871C>T (p.Arg291*) | chrX 22096976 C/T | *PHEX* | SNV | LoF, stop gained | Exon 8/22 | n.a. | n.d. | P | 372454 | #5, 6 |
|  | NM_000444.6:c.735T>G (p.Tyr245*) | chrX 22093985 T/G | *PHEX* | SNP | LoF, stop gained | Exon 7/22 | n.a. | n.d. | P | 2737109 | #4 |
|  | NM_000444.6:c.454delG (p.Asp152fs) | chrX 22077493 G/- | *PHEX* | short deletion | LoF, frameshift | Exon 5/22 | n.a. | n.d. | P* | novel | #12 |
|  | NM_000444.6:c.419C>G (p.Ser140*) | chrX 22076457 C/G | *PHEX* | SNV | LoF, stop gained | Exon 4/22 | n.a. | n.d. | P | 419416 | #18 |
|  | NC_000023.10:g.22180963_22226189del | chrX 22180963- 22226189 del | *PHEX* | LSV | Deletion | Intron 14-18 | n.a. | n.d. | n.d. | novel | #2 |
|  | NM_000444.6:c.217_218delTG (p.Val73fs) | chrX 22047081 TG/- | *PHEX* | short deletion | LoF, frameshift | Exon 3/22 | n.a. | n.d. | n.d. | novel | #8 |
| Moderate | NM_000444.6:c.1952G>C (p.Arg651Pro) | chrX 22226495 G/C | *PHEX* | SNV | Missense | Exon 19/22 | am: 0.996 (LP); CADD: 31 (MP) | n.d. | P | 438548 | #21, 30 |
|  | NM_000444.6:c.1700G>C (p.Arg567Pro) | chrX 22212958 G/C | *PHEX* | SNV | Missense, splice site | Exon 16/22 | am: 0.799 (LP); CADD: 35 (MP) | n.d. | P/LP | 372459 | #25 |
|  | NM_000444.6:c.1324G>T (p.Val442Phe) | chrX 22133544 G/T | *PHEX* | SNV | Missense | Exon 12/22 | am: 0.686 (LP); CADD: 15.87 (MB) | n.d. | P/US | 1525455 | #10 |
|  | NM_020638.4:c.526C>T (p.Arg176Trp) | chr12 4370573 G/A | *FGF23* | SNV | Missense | Exon 3/3 | am: 0.352 (A); CADD: 24.3 (I) | 6.20E-07\| n.d.\| n.d.\| n.d. | P | 1072118 | #15 |

## Discussion

In this study, the prevalence of bilateral MD-hp among male XLH patients was millions-fold higher than expected by chance in a small cohort (n=33). Most affected males had definitive bilateral MD with ES hypoplasia, whereas others exhibited a closely related severe, fluctuating/progressive SNHL phenotype combined with ES hypoplasia. In contrast, affected females showed a markedly milder endo-phenotype, typically unilateral with mild-to-moderate fluctuating/progressive SNHL without ES hypoplasia. This clear sex difference likely reflects a threshold effect of inner ear-specific *PHEX* gene dosage (Figure 5), supported by epidemiological, mechanistic, and mouse-model evidence:

**Figure 5.**
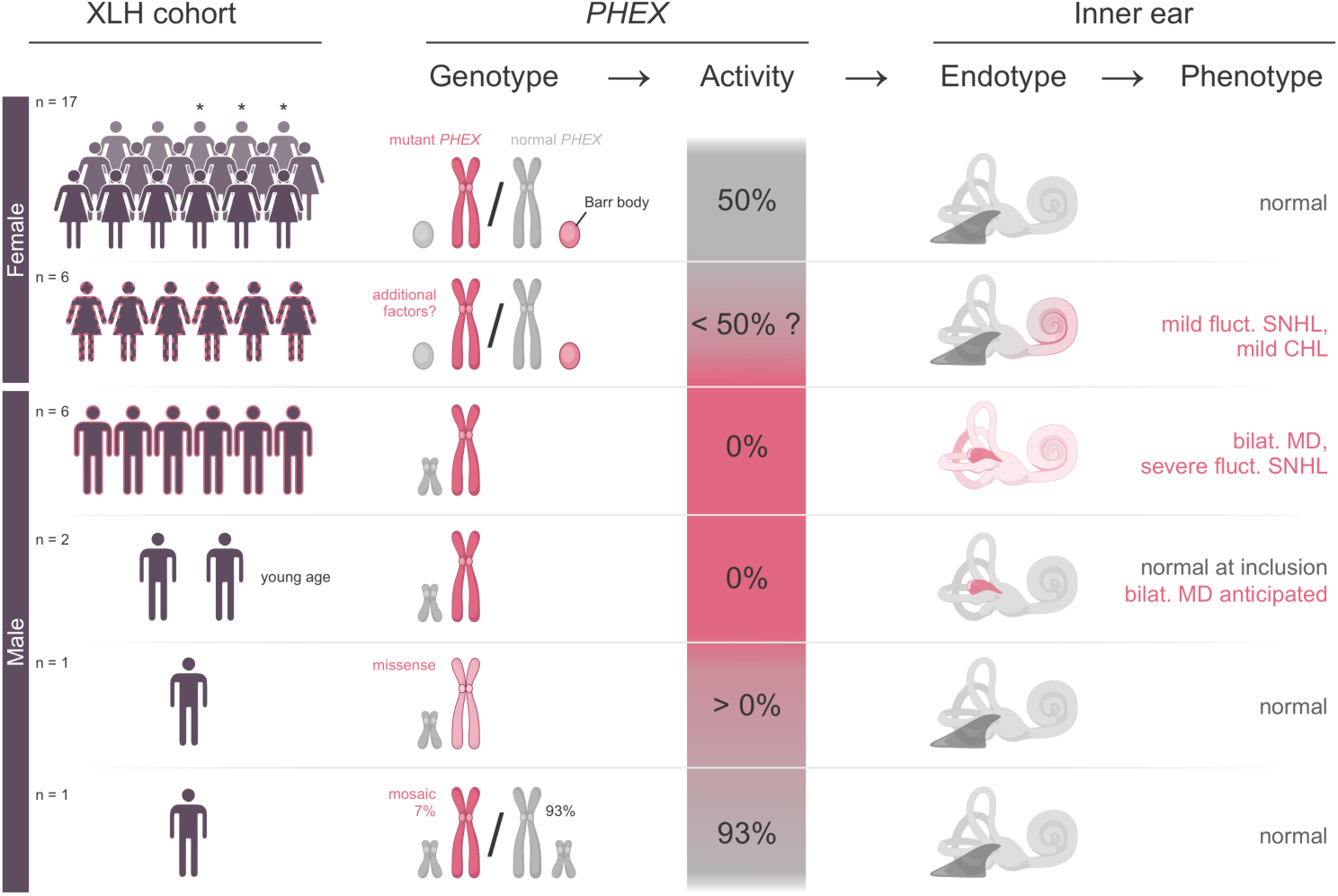
Genotype–endotype–phenotype relationships in XLH. Hemizygous males carrying a null *PHEX* allele produce no functional protein and uniformly develop the MD-hp endo-phenotype. In contrast, hemizygous males with mosaic or hypomorphic missense *PHEX* variants—and heterozygous females—retain sufficient PHEX activity, resulting in a milder or absent auditory phenotype. Asterisks (*) denote female XLH patients in whom no causal *PHEX* variant was identified.

### (i) Epidemiological evidence of a shared etiology

Our data reveal a striking epidemiological association between MD-hp and XLH: the prevalence of MD-hp among XLH patients exceeds that in the general population by six orders of magnitude. With general-population prevalences of 0.06% for MD-hp and 0.005% for XLH, the probability of their co-occurrence by chance is roughly one in 33.3 million individuals. Yet, in our combined cohorts, we identified six XLH patients with MD-hp, a finding six million-fold higher than expected by random coincidence, and thus unlikely to reflect chance alone. That all six were male, despite more female XLH carriers in our cohort, is consistent with prior reports of MD-hp predominance in males,^11,13^ and argues against a sampling bias.

### (ii) Biological plausibility of a causal relationship

Normal organ development and function require each tissue to receive a minimum level of gene activity; when expression falls below a tissue’s critical threshold, maldevelopment and disease follow. This “ultrasensitivity-mediated threshold effect”^38,39^, in which small changes in protein output can produce all-or-nothing phenotypes across different organs—depending on one organ’s threshold, explains phenotypic variability across organs in X-linked disorders—for example, complete versus partial dystrophin loss in Duchenne versus Becker muscular dystrophy breaches distinct muscle and cardiac thresholds, and skewed X-chromosome inactivation in Rett syndrome carriers can unmask severe neurologic disease despite mosaic *MECP2* expression.^40–44^ Applied to XLH, the same concept may account for the male-predominant MD-hp phenotype: hemizygous males, carrying a mutant, LoF *PHEX* allele, produce no (0%) functional protein, whereas heterozygous females, carrying a mutant LoF and a normal *PHEX* allele, retain ∼50% *PHEX* activity through random X-chromosome inactivation *PHEX* variant mosaicism. Both sexes fall below the systemic *PHEX* threshold for bone mineralization and phosphate homeostasis—explaining universal skeletal and renal manifestations—yet only males fall below the higher *PHEX* requirement of the developing inner ear, resulting in hypoplastic ES development, a pathomorphological predisposition that unfolds over decades into clinical MD-hp.

Four atypical male cases further support this ultrasensitive threshold concept: two older patients—one in their 60s with somatic mosaicism for a truncating *PHEX* mutation in ∼93% of cells, and another in their 40s carrying a hypomorphic missense variant. The deleterious effect of this missense variant has been confirmed for XLH in familial cases,^45,46^ where it segregates with the disease for bone deformities and active rickets, though no audiovestibular symptoms are indicated. This suggest that they retain enough residual *PHEX* activity to support normal inner ear development (normal ATVAs, no MD). By contrast, two younger hemizygous males (in their 20s and 30s) with full LoF mutations already exhibit bilateral ES hypoplasia (ATVA ≥LJ140°) despite lacking clinical symptoms, signaling a breach of the *PHEX* activity threshold during inner ear development that likely presages future MD-hp^12,13^.

### (iii) Dosage-dependent severity in females supports inner ear-specific PHEX threshold

Female XLH patients exhibited considerably milder audiovestibular phenotypes compared to males, including unilateral mild-to-moderate low-to-mid frequency SNHL (5 subjects), mild CHL (2 subjects), or isolated asymptomatic unilateral ES hypoplasia (2 subjects). Importantly, no significant age difference was found between female (mean 46.2 ± 17.6 years) and male patients (mean 53.1 ± 13 years), excluding age-related progression cannot explain this disparity. Although sex differences in XLH’s skeletal and renal manifestations are debated,^45,47–50^ our data strongly support an inn ear-specific, dosage-dependent effect of *PHEX*: heterozygous females retain approximately 50% PHEX activiy due to random X-chromosome inactivation and are at risk of developing a substantially milder (partial) audiovestibular phenotype—though not all manifest symptoms—compared with hemizygous males.

### (v) Towards experimental validation in Phex-deficient mice

PHEX normally cleaves and inactivates FGF23, and loss of PHEX leads to elevated FGF23 levels that drive renal phosphate wasting and the skeletal pathology of XLH. Mouse models lacking Phex faithfully recapitulate these systemic features and also develop an audiovestibular phenotype mirroring key features of human MD—with early endolymphatic hydrops, progressive cochleo-vestibular degeneration, and SNHL.^37,51,52^ Our preliminary data indicate that *Phex*-null males display more severe inner ear pathology than females, mirroring the sex- and X-chromosome dosage effects we observe in XLH patients (unpublished data). Ongoing studies will test whether this inner ear phenotype stems from dysregulated FGF23 signaling and phosphate balance, altered vitamin D metabolism, or another *Phex*-dependent pathway. If these results hold up, they would provide crucial support for our inner ear dosage-threshold model and establish *Phex* deficiency as the causal, male-predominant driver of MD in XLH.

### Limitations

Our cohort’s modest size (n = 33) and the rarity of XLH limit generalizability, although our epidemiological signal and consistency with prior male-biased MD-hp reports are compelling. The etiology of CHL observed in two female patients remains unclear, though ossicular chain hypomineralization has proposed as a potential mechanism in both XLH humans and *Phex*-deficient mouse models.^53^ We cannot yet exclude additional modifiers—such as sex hormones, skewed X-chromosome inactivation, epigenetics, or modifier genes—that may influence inner-ear susceptibility. Outlier cases in both sexes highlight the complex genetic and epigenetic landscape governing MD-hp risk and underscore the need for larger, prospective XLH cohorts to refine genotype-phenotype correlations and personalized risk stratification.

### Clinical impact and future directions

Our study is the first to link a clearly defined *PHEX* genotype with an inner ear endotype and the clinical MD phenotype, establishing that hemizygous males with pathogenic *PHEX* variants are at uniquely high risk for bilateral MD. By combining male sex, early diagnosis of XLH diagnosis, and abnormal ATVA measurements (detectable from early adulthood), clinicians can now identify individuals likely to develop MD-hp years—if not decades—before symptom onset. This predictive capacity opens the door to personalized counseling, early monitoring, and the possibility of preventive interventions to preserve inner-ear function. Mechanistically, future work should employ *Phex*-deficient mouse models and in vitro systems to dissect *PHEX*’s role in endolymphatic sac development, define its downstream MD-driving pathways, and screen candidate therapies. The diversity of *PHEX* variants in our cohort— hemizygous male null alleles (complete LoF), heterozygous female null alleles (∼50% function), missense variants (presumed partial LoF), and mosaicism—underscores the need for deeper genetic analyses (e.g., multi-tissue sampling to quantify mosaic burden; functional assays of missense alleles) to correlate genotype with inner-ear pathology. Such translational efforts promise not only to validate our tissue-specific functional-threshold framework but also to pave the way for targeted strategies that prevent or delay MD-hp in XLH patients.

## Data Availability

All data produced in the present study are available upon reasonable request to the authors

## Acknowledgements

We thank Dr. Thomas Carpenter and Dr. Karl Insogna for their help with patient recruitment. This work was supported by the Hearing Health Foundation, the Dorothy Wolff Fellowship in Otolaryngology Research, and the Herbert Silverstein Grant.

